# An automatic diagnostic system for pediatric genetic disorders by linking genotype and phenotype information

**DOI:** 10.1101/2021.08.26.21261185

**Authors:** Xinran Dong, Bingbing Wu, Huijun Wang, Lin Yang, Xiang Chen, Qi Ni, Yaqiong Wang, Bo Liu, Yulan Lu, Wenhao Zhou

**Affiliations:** Molecular Medical Center, Children’s Hospital of Fudan University, Shanghai, 201102, China; Key Laboratory of Birth Defects, Children’s Hospital of Fudan University, Shanghai, 201102, China

**Keywords:** diagnostic system, genetic disorders, genotype, phenotype

## Abstract

**Background:** Quantitatively describe the phenotype spectrum of pediatric disorders has remarkable power to assist genetic diagnosis. Here, we developed a matrix which provide this quantitative description of genomic-phenotypic association and constructed an automatic system to assist the diagnose of pediatric genetic disorders.

**Results:** 20,580 patients with genetic diagnostic conclusions from the Children’s Hospital of Fudan University during 2015 to 2019 were reviewed. Based on that, a phenotype spectrum matrix -- cGPS (clinical Gene’s Preferential Synopsis) -- was designed by Naïve Bayes model to quantitatively describe genes’ contribution to clinical phenotype categories. Further, for patients who have both genomic and phenotype data, we designed a ConsistencyScore based on cGPS. ConsistencyScore aimed to figure out genes that were more likely to be the genetic causal of the patient’s phenotype and to prioritize the causal gene among all candidates. When using the ConsistencyScore in each sample to predict the causal gene for patients, the AUC could reach 0.975 for ROC (95% CI 0.972-0.976 and 0.575 for precision-recall curve (95% CI 0.541-0.604). Further, the performance of ConsistencyScore was evaluated on another cohort with 2,323 patients, which could rank the causal gene of the patient as the first for 75.00% (95% CI 70.95%-79.07%) of the 296 positively genetic diagnosed patients. The causal gene of 97.64% (95% CI 95.95%-99.32%) patients could be ranked within top 10 by ConsistencyScore, which is much higher than existing algorithms (p <0.001).

**Conclusions:** cGPS and ConsistencyScore offer useful tools to prioritize disease-causing genes for pediatric disorders and show great potential in clinical applications.

## Introduction

The rapid and accurate identification of disease-causing genes is particular important to clinical geneticists, which could facilitate clinical decision-making, improve prognosis, and help with further family planning[1]. Exome-based NGS (next-generation sequencing), including WES (whole-exome sequencing) and CES (clinical exome sequencing), are cost-effective options for detecting candidate disease-causing variants in genetic disorder patients[2-4]. Meanwhile, databases including OMIM[5], Orphanet[6], and human phenotype ontology (HPO)[7] have collected and summarized clinical phenotype for genetic diseases from previous studies, offering reliable phenotype reference for patient genetic diagnosis. However, these genomic-phenotypic recordings are rarely quantified (or weighted), making it difficult to be automatically applied in the priorization of disease-causing gene.

Currently, many algorithms have helped to calculate the association between genotype and phenotype. Phenolyzer [8], PhenoPro[9, 10], and Phen2Gene[11] focus on obtaining candidate gene lists by using clinical phenotypes to query database-derived information (such as OMIM and Ophanet); Exomiser (phenix, hiPHIVE)[12], Xrare[13] and Phen-Gen[14] were designed to prioritize genes and variants by estimating the variant’s pathogenicity and the consistency between patient’s phenotype with public reported. However, most of these tools were evaluated on simulation dataset or small-scale public dataset and cannot reliably identify causal genes in real clinical dataset[15].

The Children’s Hospital of Fudan University has been involving in clinical genetic tests with NGS since 2013, where the genetic diagnosis was made by experienced clinicians and genetic counselors. Several studies of disease cohort focusing on pediatric genetic disorders have been previously described and published[16, 17], sharing valuable experience and discoveries in the diagnosis of pediatric genetic disorder. These large-scale clinical findings provide huge resources for calculating the phenotype spectrum of pediatric genetic disorders and developing tools to assist genetic interpretation. In this study, we systematically summarized the genotype, phenotype and genetic diagnosis conclusion information of 20,580 patients, of which 3,507 were positive diagnoses. Based on this real large-scale cohort, we developed a matrix named cGPS (clinical Gene’s Preferential Synopsis), which provided a quantitative description of the clinical phenotypes that may be affected by each known disease-causing gene. Based on cGPS, we further provided an intuitive artificial intelligence system, “ConsistencyScore”, which can successfully prioritize genes that might cause the patient’s clinical phenotype. In another independent validation cohort of 2,323 patients with 296 positive genetic diagnosis, ConsistencyScore could rank the causal gene as first for 75.00% of the patients, and in the top 10 for 97.64%. In general, cGPS matrix and ConsistencyScore could be great resource and tool for clinical genetics community.

## Results

### 1. Cohort for the automatic diagnostic system

In total 23,251 patients from the CHFU were recruited, together with their genotypic, phenotypic and genetic-diagnostic information (**Figure 1A)**. Based on these genetic diagnostic conclusions, we designed the cGPS matrix to quantify the spectrum of affected clinical categories when a certain gene was damaged by variants. With cGPS to describe genotype-phenotype associations, we further provided and tested its first clinical application – a patient-specific “ConsistencyScore” -- to automatically prioritize disease-causing genes for genetic disorder patients with hundreds of genes affected by variants identified with NGS **(Figure 1B)**.

**Figure 1.**
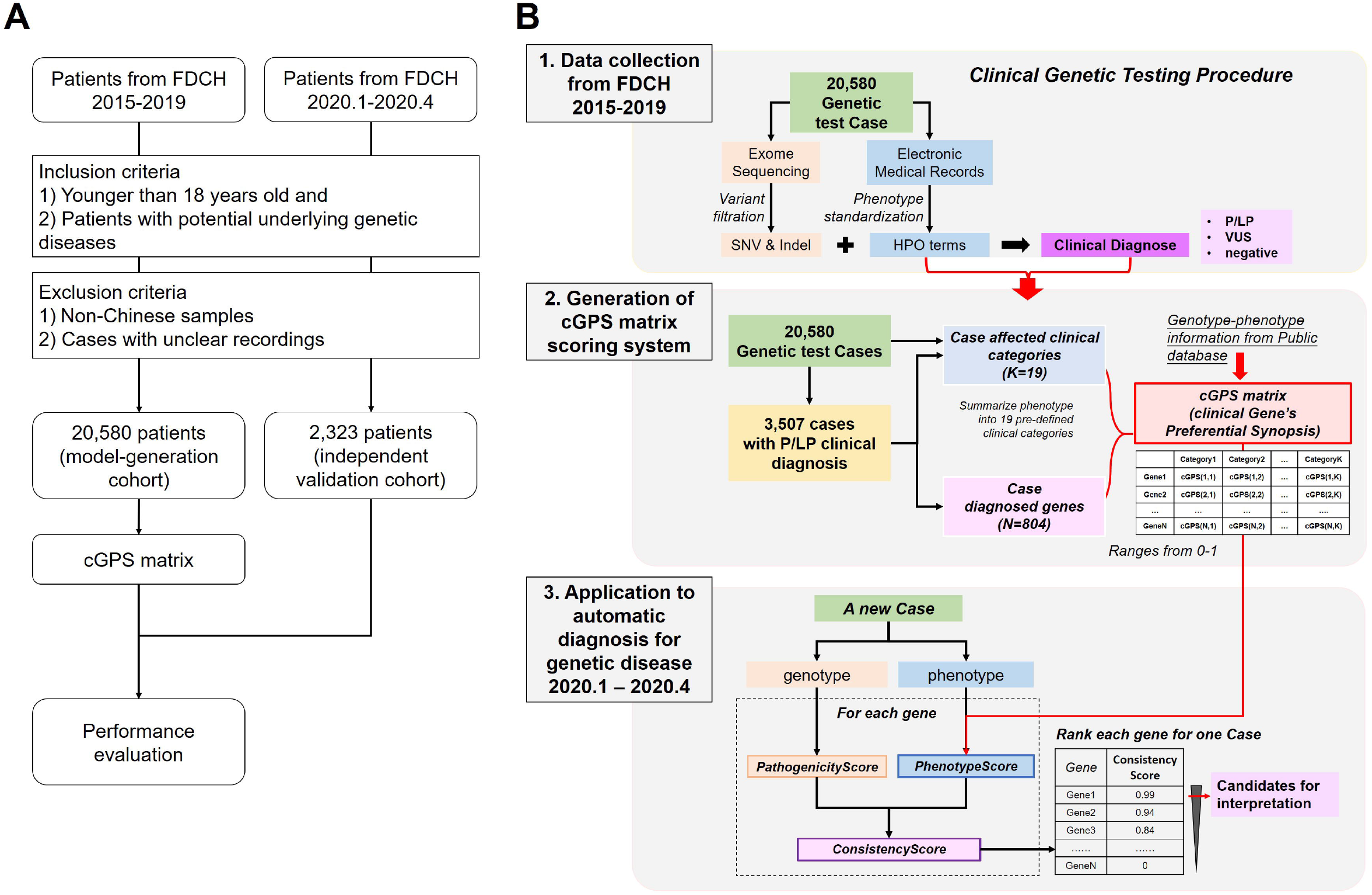
Flow chart for (A) study design and (B) predictive model design. (A) In this study for developing and validating a predictive model to prioritize disease-causing genes, two cohorts were recruited, for model-generation and independent validation. (B) The generation of predictive model. Data collection: Variants analysis with exome sequencing was performed and clinical information was automatically extracted into HPO terms with manual review. Clinical diagnosis was made by genetic counselors on candidate list after automatic variant filtration step. 20,580 genetic testing cases were collected from FDCH from 2015-2019, from which 3,507 cases were with P/LP clinical diagnosis, in total 804 genes have been ever P/LP reported. The HPO terms were further summarized into clinical categories. This information was collected from one center with unified standard and were further summarized into genotype, phenotype and diagnose-level. Generation of cGPS matrix: The cGPS matrix score was calculated from all P/LP-diagnosed samples in CCGT dataset and public database (OMIM and geneOrganizer). The score ranges from 0-1 and higher scores indicate higher correlation between the gene and the clinical category. Application to automatic diagnoses for genetic disease: cGPS matrix could further assist the automatic prioritizing of disease-causing genes by calculating PhenotypeScore for each candidate gene. The final ConsistencyScore that incorporate PhenotypeScore and PathogenicityScore could be used for disease-causing gene’s priorization, and higher score genes could have higher probability of disease-causing. Detailed procedure could be found in Methods. Abbreviation. P/LP: pathogenic/likely pathogenic. VUS: variant of unknown significance.

For the model generation cohort with 20,580 patients, 15,622 patients were tested by clinical exome sequencing (CES) and 4,958 by whole-exome sequencing (WES) (**Table1)**. There were in total 12,583 males and 7,997 females, 79.7% of them were smaller than five years-old **(Supplementary Figure1A)**. A total of 3,507 patients (17.0%) had a positive genetic diagnosis result, which were referred to as genetic conclusions including P/LP-level variants. The positive genetic results involved 3,678 variants from 804 genes. Top diagnosed disease-causing genes were *ATP7B* (n=343), *SLC25A13* (n=121), *NF1* (n=120), *DMD* (n=109) and *SCN1A* (n=108), consist 21.7% of all diagnosed patients **(Supplementary Figure1B)**.

**Table1.**
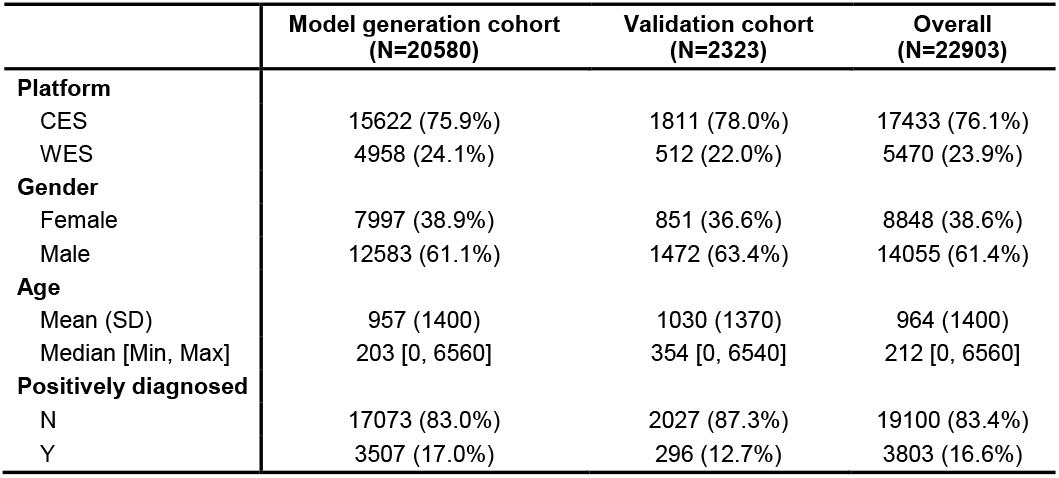
Characteristics of patients in the model generation and validation cohort.

Top three HPO terms were HP:0000952 (Jaundice, 16.3%), HP:0001250 (Seizure, 15.8%) and HP:0002904 (Hyperbilirubinemia, 15.2%) **(Supplementary Figure1C)**. If summarize original HPO terms into root HPO terms under HP:0000118 (Phenotype abnormality), top three root HPO terms were HP:0000707 (Abnormality of the nervous system, 41.2%), HP:0025031 (Abnormality of the digestive system, 36.7%) and HP:0001939 (Abnormality of metabolism/homeostasis, 35.7%) **(Supplementary Figure1D)**. For simplicity in both research and application, and by integrating original HPO terms with HPO root terms, patient’s clinical phenotypes were summarized into 19 pre-defined clinical categories (**Supplementary Table 1**). The largest clinical category was nervous system (NV) that included 45.4% of the patients, followed by digestive system (DG, 36.6%), metabolism (MB, 35.3%) and immune system (IM, 29.1%). One patient could be affected by phenotypes belongs to multiple clinical categories simultaneously. Overall, several clinical category pairs were highly overlapping (or often appear simultaneously in patient) such as digestive system and liver (DG and LV, 75.4%), jaundice and liver (JD and LV, 59.0%) and jaundice and skin (JD and SK, 51.7%) (**Supplementary Figure2**).

### 2. Generation of cGPS matrix

The cGPS matrix is a knowledge-based score matrix which describes the probability to cause the phenotype of a clinical category when a certain gene is affected by genetic variants (See Methods for details). We could only use information from CCGT dataset or integrate with public databases (OMIM and GeneOrganizer) to generate cGPS matrix. Meanwhile, three phenotype manipulation strategies were considered, i.e based on original HPO terms, root HPO terms and 19 pre-defined clinical categories. Only 804 genes could get cGPS matrix if only use CCGT dataset and for those genes, when integrating public databases, the scores were highly consistent (all *P*<2e-16, Spearman Correlation Coefficient 0.76, 0.72, 0.53). Secondly, mean/median/max cGPS matrix score for genes between using 19 pre-defined clinical categories and HPO root terms, original HPO terms, were significantly highly consistent (all *P*<2e-16, Spearman Correlation Coefficient>0.7) but significantly higher score values were detected in the pre-defined clinical categories (all *P*<2e-16). Taking together, cGPS matrix based on pre-defined clinical categories showed as more robust with the integration of public databases and can bring better score discrimination for subsequent applications.

In total 608 genes from cGPS matrix based on pre-defined clinical categories had maximum score higher than 0.5. Clinical categories that tend to co-occurrent also have similar cGPS pattern, such as jaundice (JD) & liver (LV), digestive system (DG) & metabolism (MB) (**Supplementary Figure3A**, categories were clustered on the left panel). cGPS score pattern for NV (nervous system) were different from all other categories, and it include the most related causal genes (359 genes with cGPS score > 0.5) and highly associated genes (147 genes with cGPS score > 0.7), followed by clinical categories such as DG (digestive), MB (metabolism), and LV (liver) **(Supplementary Figure3B)**.

### 3. Diagnostic performance of the automatic diagnostic system

To apply cGPS in genetic diagnosis, we designed three scores on patient-gene pair level, i.e PathogenicityScore, PhenotypeScore and ConsistencyScore (See Methods for details). Since genes in each patient could be labeled as P/LP, VUS or B/LB, we evaluated the score distribution of the patient-gene pairs under different labeling results. For patient-gene pairs labeled as P/LP, all three scores were significantly higher than other labels (all *P*<2e-16). However, for patient-gene pairs with PathogenicityScore higher than 0.9, only 2.53% of them were reported as P/LP, but for PhenotypeScore and ConsistencyScore the percentages were 19.7% and 80.2%, separately **(Supplementary Figure4)**.

We further tested the performance of ConsistencyScore **(Figure 3A)**. Firstly, ConsistencyScore can achieve good performance in predicting P/LP-diagnosed sample-gene pairs in 6,174 testing dataset (AUC 0.965 for ROC with 95% CI 0.960-0.969, and AUC 0.281 for precision-recall curve with 95% CI 0.255-0.311) **(Figure 3B-C)**. Secondly, the gene’s rank ordered by ConsistencyScore in each sample could get better performance in predicting P/LP-diagnosed gene in 1,005 P/LP-diagnosed testing dataset (AUC 0.975 for ROC with 95% CI 0.972-0.976, and AUC 0.575 for precision-recall curve with 95% CI 0.541-0.604) **(Figure 3D-E)**. Meanwhile, 69.0% could rank 1 and 96.1% rank within top 10 **(Figure 3F)**.

**Figure 2.**
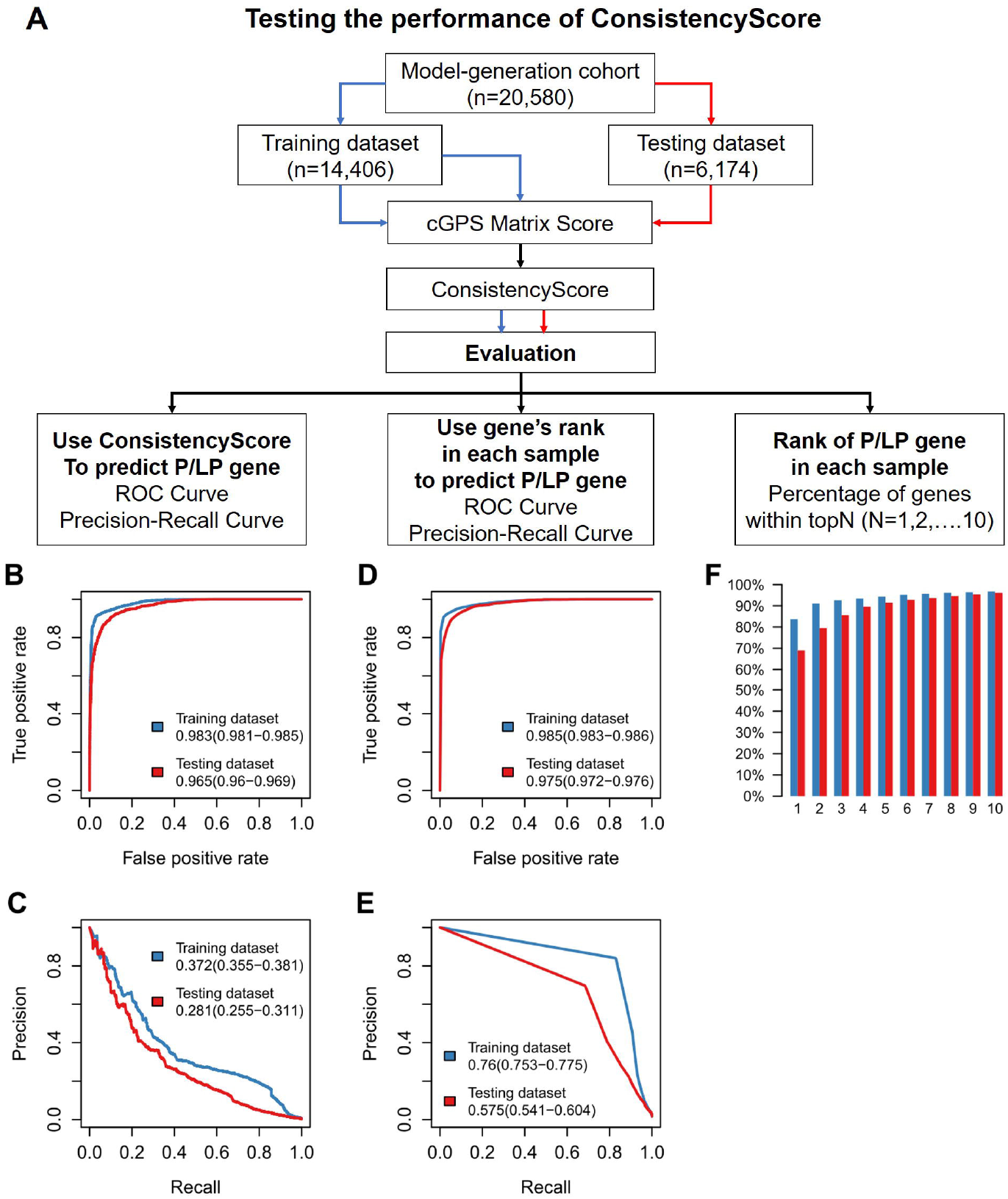
Performance ConsistencyScore in predicting P/LP-diagnosed genes in training and testing dataset. (A-B) Testing the performance of ConsistencyScore to predict diagnostic sample-gene pairs in training and testing dataset. (C-D) Testing the performance of gene’s rank ordered by ConsistencyScore in each sample to predict diagnostic sample-gene pairs in training and testing dataset. ROC curves (A,C) and precision-recall curves (B,D) were used to evaluate the performance. AUC values with 95% CI were shown in each sub-figure. (E) The percentage of cases with final diagnostic-genes within top candidate number (1,2…10) in training dataset (blue) and testing dataset (red). The cGPS matrix was calculated by using only training dataset with original HPO terms summarized into 19 pre-defined clinical categories. In A-B, all samples were used for evaluation. In C-E, only samples with P/LP-diagnostic genes were used for evaluation.

**Figure 3.**
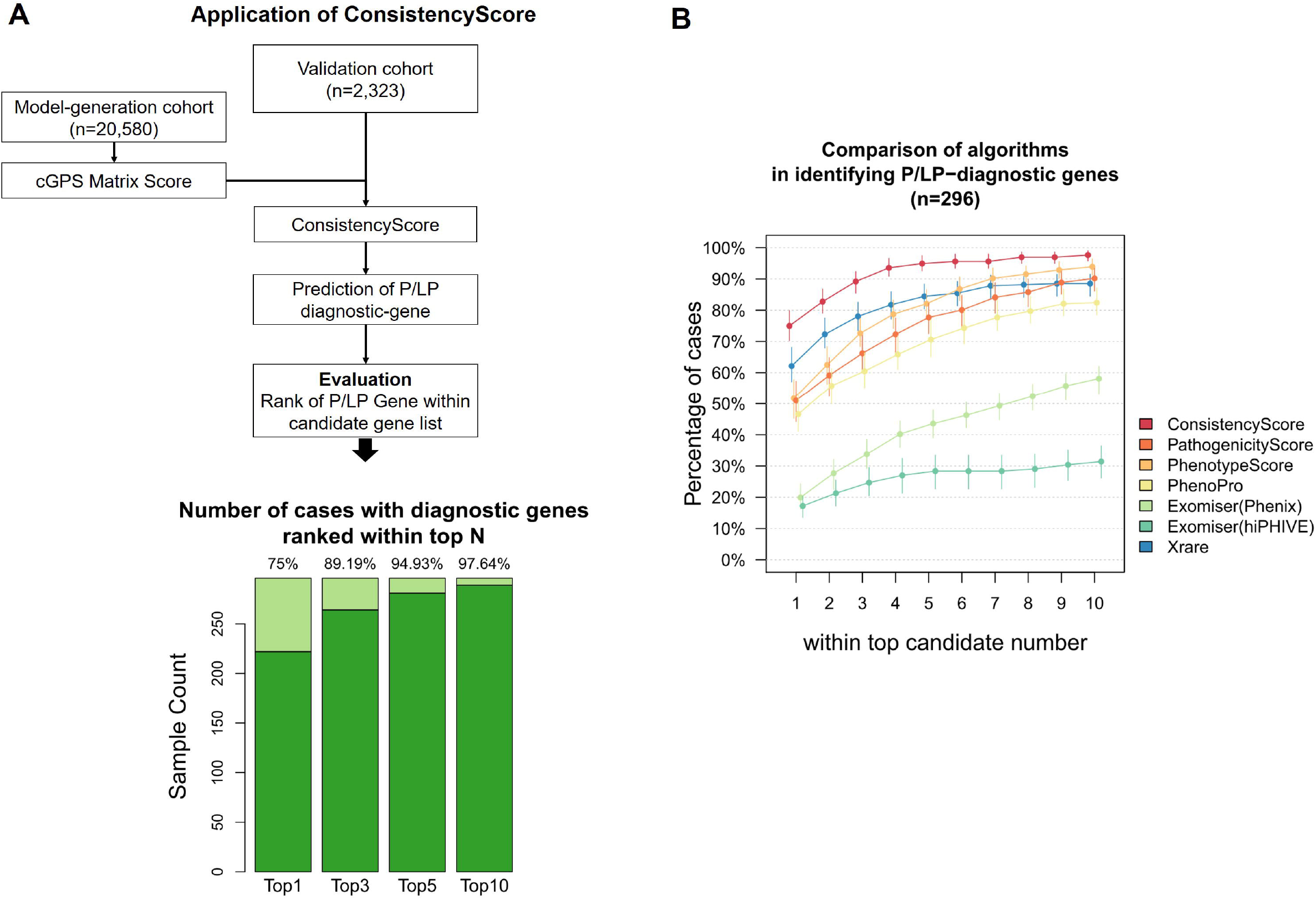
Performance evaluation ConsistencyScore in priorization disease-causing genes from an independent dataset with 296 P/LP-diagnosed patients. (A) Application of ConsistencyScore in a new independent dataset. Count of samples and percentage of the reported P/LP-diagnosed genes within top 1,3,5,10 of each sample’s candidate variant list were shown with 95% CI. In 296 samples, the 75% manually reported P/LP-diagnosed genes had the highest ConsistencyScore value. (B) The percentage of cases with final diagnostic-genes within top candidate number (1,2…10) produced by each algorithm were shown, with 95% CI bar. Compared with other tools, ConsistencyScore could get the best performance.

In addition, the performance of ConsistencyScore to predict P/LP-diagnosed sample-gene pairs using pre-defined clinical categories were significantly higher than using HPO root terms (AUC 0.963 for ROC with 95% CI 0.959-0.967, *P*=0.02) and original HPO terms (AUC 0.956 for ROC with 95% CI 0.949-0.960, *P*=1e-4) (**Supplementary Figure 5A**,**B**,**F**,**G**). Similar performance were shown for using gene’s rank ordered by ConsistencyScore in each sample compared with using HPO root terms (AUC 0.974 for ROC with 95% CI 0.971-0.975, *P*=0.3) and but higher than using original HPO terms (AUC 0.964 for ROC with 95% CI 0.961-0.967, *P*=3e-7) (**Supplementary Figure 5C**,**D**,**H**,**I**). Besides, the performance of PathogenicityScore and PhenotypeScore to predict P/LP-diagnosed genes were all significantly lower than ConsistencyScore in all comparisons (all *P*<2e-16) (**Supplementary Figure 6**). Specially for the precision-recall curve, the PathogenicityScore and PhenotypeScore were particularly worse, demonstrating their insufficient ability to make true P/LP predictions.

### 4. Performance of ConsistencyScore in MGHA corpus and local new independent validation dataset

The Melbourne Genomics Health Alliance (MGHA) variant prioritization framework contains 47 samples with phenotype (HPO terms) and reported variant information[18]. We calculated ConsistencyScore for each reported gene (in total 59 genes) with estimated P/LP probability and not P/LP probability (**Supplementary Figure 7A**). In total 41 samples (53 genes) had great P/LP predictions (marked as P) and 5 samples (5 genes) showed not significantly difference of ConsistencyScore between P/LP-reported and not P/LP-reported in CCGT dataset (**Supplementary Figure 7B**). Only one samples failed the prediction, i.e 0204339 with *NOTCH2*. The patient was recorded to be affected with abnormalities in face and bone. However, the cGPS score for *NOTCH2* (MIM:610205) is high in DG (digestive), LV (liver), JD (jaundice), though had score but low in SL (skeletal), MX (maxillofacial) and PD (polydactyly/abnormal of limbs). Lack of typical phenotype observed failed good P/LP prediction.

To further show how the ConsistencyScore would help genetic diagnosis in real clinical practice, we further collected 2,323 patients (including 296 P/LP diagnosis, **Figure 4)**. ConsistencyScore for P/LP patient-gene pairs from patients with P/LP conclusion (positive cases) were significantly higher than other patient-gene pairs in these positive cases (i.e., in patients with positive conclusions, genes labeled as P/LP compared with other labels), and higher than all patient-gene pairs in cases with no P/LP conclusions (*P*<2e-16). No significantly difference was observed between the latter two groups. In 75.00% (95% CI 70.95%-79.07%) of the patients with positive conclusion (described as P/LP-diagnosed patients), the ConsistencyScore could rank the P/LP genes as the first among other genes in that patients (defined as the ranked-first percentage in the following part). And in 97.64% (95% CI 95.95%-99.32%) of these P/LP-diagnosed patients, the ConsistencyScore could rank the P/LP genes within top 10 (**Figure 4)**.

From 296 P/LP-diagnosed patients, 268 of them were diagnosed with genes from the previous 804 P/LP genes in the model-generation cohort, i.e their PhenotypeScore and ConsistencyScore based cGPS matrix was mainly relied on observations provided in this study. The 28-remaining diagnostic-genes were never labeled as P/LP in genetic reports of our cohort and their cGPS matrix is mainly based on genotype-phenotype recordings from public databases. Here, the ranked-first percentage for these 268 cases could reach 80.22% (95% CI 75.27%-85.43%), and was significantly higher than the 28 not reported cases (*P*<2e-16, **Supplementary Figure8A**). The difference demonstrated that the cGPS matrix originated from real P/LP observations do contribute greatly for the diagnosis of new cases. Besides, ConsistencyScore shows significant greater performance for CES platform than WES (ranked-first percentage 80.61% vs 67.93%, *P* = 6e-3) (**Supplementary Figure8B**). Disease-causing genes with AD-inherited model (ranked-first percentage 68.15%) shows worse performance than AR (81.36%, *P* = 1.1e-4) and X-linked (79.07%, *P* = 5.5e-3, **Supplementary Figure8C**). Generally, the performance of ConsistencyScore was relatively robust in different scenarios, except for diagnosing genes that were not reported as P/LP before.

Further, we compared the result of Exomiser(Phenix), Exomiser (hiPHIVE), Xrare and our previously published algorithm PhenPro on these 296 P/LP-diagnosed patients with default parameters. Compared with other tools, ConsistencyScore shows the best performance in identifying P/LP-diagnostic genes (**Figure 4B**). For the two scores only rely on phenotype information, PhenotypeScore had better performance than PhenoPro (ranked-first percentage 51.69% v.s. 46.62%, *P*<2e-16). Xrare takes the pathogenicity potential of variants into consideration and thus performed better ranked-first percentage (62.16%) than the previous two methods, but it also made 25 false negative cases of which the disease-causing genes were absent in the final list. When checking different conditions, Exomiser (Phenix, hiPHIVE) shows relatively poor performance (about 20% ranked-first percentage) with default parameters and is worse for AR-inherited genes (4.23% ranked-first percentage, mainly because the inheritance model is not used in Exomiser) (**Supplementary Figure8I**). Besides, all tools had better performance for X-linked genes (**Supplementary Figure8J**). In general, ConsistencyScore has demonstrated its great power in assisting the disease-causing gene prioritization.

## Discussion

Pediatric genetic disorders, such as congenital anomalies, are the leading cause of childhood death with age under five, especially for infants[19]. Genetic diagnosis of these disorders would help the clinical treatment and prognosis. Making genetic diagnosis requires precise and efficient identification of pathogenic variants in disease-causing genes. Currently the ability to identify variants is not a limitation, but the data interpretation is still a challenge. After accumulating a large amount of data, we could now summarize the experience in genetic diagnosis into an automatic tool, and share with our colleagues especially for the diagnosis of pediatric genetic disorders.

ACMG has provided guideline for the interpretation of variants for a certain patient with genetic disorders. Two main aspects are essential: whether the variant is pathogenic to damage the function of a certain gene or to gain new disturbing function that would cause disease, and whether the patient’s clinical phenotypes could match the expected symptom of the disorder that was caused by this gene. Unlike other phenotype-based tools discussed in this study, cGPS is a genetic-driven assessment that “predict” the possible phenotypes for patient with genetic testing. Based on the “prediction” or “expectation” of the clinical phenotype, the genetic counselor could decide whether a variant is the causal. Clinicians could effectively make follow-up plans to focus on the occurrence and development of these clinical phenotypes, and update the genetic diagnosis report when new phenotypes appear.

Here, cGPS calculation process is independent to clinical categories’ design, and in this study, we tried one pre-defined clinical categories, which showed better performance than original HPO terms and HPO root terms. Most pre-defined categories could directly match the settings of clinical department, while others (such as jaundice) are common clinical phenotypes. However, designed categories are intersected to each other, which may be biased in ConsistencyScore calculation. We checked the performance of P/LP-diagnosed gene’s rank ordered by ConsistencyScore in each sample in the testing dataset and found no significant difference of performance found between any pair of clinical categories (in total 19*18/2=171 pairs, all adjusted *P*>0.1). Besides, in the genetic screening scenario, cGPS matrix could act as a reference to design sequencing panels with specified clinical phenotype category. In cGPS calculation, not only P/LP-diagnosed samples were used but all samples were useful in estimating background phenotype distribution.

The value of ConsistencyScore could get good discrimination of P/LP-diagnosed genes to VUS and negative ones but within one sample, we strongly suggest to consider gene’s rank ordered by ConsistencyScore first to avoid possible false negative predictions. The AUC for ROC by using ConsistencyScore to predict sample-gene pairs only in 1,005 P/LP-diagnosed testing dataset (AUC 0.964 for ROC with 95% CI 0.959-0.967) was nearly the same by using all testing samples in Figure 3A, but was significantly lower than using gene’s rank in Figure 3C (*P*=2.2e-13), which demonstrated better choice of gene’s rank in practice. However, in the public validation MGHA corpus, no original genotype information available and we could only calculate ConsistencyScore for the reported genes and estimate whether the value could be a good P/LP prediction. The results showed that 53/59 (89.8%) of the P/LP-reported genes could get higher P/LP probability than not (OR>2), partially demonstrating the reliability of ConsistencyScore.

Meanwhile, there are already many tools for prioritizing disease-causing genes from patients’ sequencing data. Among them, Exomiser is one of the earliest and Xrare is one of the latest software. The calculation of PhenotypeScore is inspired from algorithms that calculate patient similarity to diagnosis new case. Here, cGPS could act as a summary of previous patient’s phenotype profile. In this study we also compared our previously published algorithm PhenoPro. Instead of using simulated dataset (which was used to evaluate these algorithms in their own research), in this study we tested them in a large-scale clinical cohort which could reflect the complicated clinical conditions and provide more objective results for clinical application. The ConsistencyScore show the best performance among different scenarios and even the PhenotypeScore could outperforms PhenoPro. Xrare missed the disease-causing gene for 25 patients, including *G6PD* (in 5 patients) and *GJB2* (in 4 patients). The allele frequency of these missed variants are around 0.3%∼0.5% in East Asian[20]. That might be the missed reason because Xrare applied population allele frequencies as features in the model. As a result, pathogenic variants with relatively higher local allele frequency might be missed. Finally, cGPS and ConsistencyScore were straightforwardly designed, transparent in the application process, requires little calculation (finish in seconds) and could easily be incorporated into other pipelines. By comparison, Exomiser took about one minute and Xrare took 5.3 minutes per sample on average.

### Limitations and future directions

Firstly, in the PathogenicityScore designing process, the treatment for protein truncation variants is relatively simple. We are trying to include strategies to predict nonsense-mediated mRNA decay for improvement. Besides, the choice for missense variants was currently based on REVEL and more scores such as CADD could also be tried. Secondly, the performance of ConsistencyScore in the testing dataset is lower than the validation dataset, which is because of 33% percent more samples included in the cGPS matrix generation. The performance increased slowly with more samples included but how many samples are sufficient enough to get a robust cGPS matrix need to be further explored. Thirdly, for patients that ConsistencyScore was failed to prioritize the P/LP genes, in only 7 patients the P/LP genes were out of top 10. Here we selected one for detailed discussion. This patient was diagnosed as Leukodystrophy, hypomyelinating, 7 [MIM: 607694] caused by variant on *POLR3A*. However, in the model-generation dataset no patients were P/LP-diagnosed as *POLR3A* and the cGPS was calculated by referring phenotype recordings from public databases, making this gene rank lower than others. Updating cGPS with more P/LP-diagnosed patients will improve the performance. Finally, how to quantify the pathogenicity of CNV is still a future direction.

## Conclusions

The findings of this study suggest that an automatic diagnostic system based on quantifying phenotype spectrum can be used as a prospective tool for disease-causing gene’s prioritization in pediatric disorders.

## Methods

### 1. Participants and source of the data

This study developed an automatic diagnostic system for pediatric genetic disorders. We generated a predictive model based on a retrospective cohort with 20,580 patients (from 2015 to 2019) from the Children’s Hospital of Fudan University (CHFU). The model was further validated in an independent cohort of another 2,323 patients (from January 2020 to April 2020) from the CHFU. Patients underwent genetic tests were recruited, dominated by the Chinese Han population (>99.9%). Patients were enrolled based on the following criteria: 1) age under 18 years old and 2) was suspected of genetic disorders by his/her physician. Informed consent was obtained from the parents of each patient. Genetic counselling was performed by physicians prior to the testing. Each patient would receive a genetic diagnostic report with confirmed genetic diagnose conclusions. In this study, the latest version of information (genotype, phenotype and genetic diagnose) was used if any update exists. The patients’ age was represented by the age at the time of the genetic testing. The clinician reviewed the clinical record and would exclude the patient if the phenotype record was not clear.

### 2. Genetic testing in Children’s Hospital of Fudan University

#### 2.1 Sequencing, variant calling and pre-filtering

DNA libraries of CES used the Agilent SureSelectXT Human ClearSeq Inherited Disease Kit and WES used Agilent SureSelectXT Human All Exon V5 Kit, respectively. Sequencing was performed on the Illumina HiSeq 2500 with 125 bp (2015-2016.3) or X10 with 150 bp (2016.3-2020) pair-end sequencing. The basic quality statistics were shown in **Supplementary Table 1**.

Data analysis pipelines were published in our previous studies [16, 17]. In this study, only SNVs and small insertions and deletions (InDels) were included. Briefly, raw reads were mapped to hg19 with BWA [21]and further processed by GATK3 to obtain SNVs and InDels for each sample. Except for those annotated in HGMD[22], ClinVar[23] and our internal curated databases, variants were filtered by exon region extended by 15 bp. Allele frequencies from 1KG (1000g2015aug), ExAC (exac03nontcga) and gnomAD (gnomad211_genome) were downloaded from the ANNOVAR database and annotated to variants. Next, variants with high homozygous reporting counts and high allele frequencies from the 1000 Genome (1KG), ExAC, gnomAD and internal databases were removed. The remaining variants will be used to calculate internal allele frequency and further annotated by ANNOVAR[24], VEP[25], and inheritance pattern if parental samples were available. Further filtration steps were detailed described in previous publication [16], including the 1) genes not in the black list; 2) the zygosity for the variant does not fit the gene’s inheritance model, e.g heterozygous variants in AR-inherited genes; 3) for family-based samples, the variants are not homozygous in parents; or for AD-inherited genes, the variant does not inherited from parental samples; 4) low quality or out-side capture region or high allele frequency variants not in the white list. The white list includes all public reported pathogenic variants. This step could leave approximately dozen genes (median 40) per-sample for further manually review.

#### 2.2 Phenotype processing procedure

The HPO terms for each patient sample from 2015-2016 were curated manually by clinicians. From 2017, a semiautomatic system was applied to extract HPO terms from the electronic medical record. The core of this system was the local sematic database. The English version was initially from HPO database (start from version 2016-04-01, update each year), and the Chinese version was initially from CHPO database (start from version 2016-03, update if CHPO updated). If the phrase cannot match the existing semantic database, the clinical phenotypes in Chinese were translated into English, transferred into UMLS standard phrases by MetaMap [26] and converted to HPO terms [7]. Then, the new matching items would be added into database after manually curation. In total 574 new phrases were added till now. When applying the system, all queried HPO terms would be curated by clinicians before assigning to the patient sample.

In the following cGPS matrix calculation, the HPO terms could be directly used or summarized into predefined groups. Three strategies were tested in this study. The first one was to use the original HPO terms. The second one was to summarized the HPO terms into their root terms (the first level child nodes under HP:0000118: Phenotype abnormality). The third one was to summarize the HPO terms into 19 predefined clinical categories. The category settings were based on clinical departments and the frequency of phenotypes. A detailed description of the 19 clinical categories and related HPO terms can be found in **Supplementary Table 2**. For each HPO term listed in this relationship table, all recursive child terms would also be included. Some HPO terms could belong to multiple categories (e.g., HP:0002090, Pneumonia, belongs to both RP and IM), and some HPO terms could not match any of the clinical categories (e.g., HP:0001622, Premature birth). For those 20,580 patients, we obtained a 19-column binary table indicating the affected clinical category.

#### 2.3 Genetic diagnose procedure

The standard for variant classification as P/LP (pathogenic/likely pathogenic) was based on the ACMG guidelines but with some adjustment, described in our previous work[17]. Specifically, 1) the variant would likely explain the indication for testing and may be responsible for the patient’s clinical presentation, and 2) the variant has the same amino acid change as a previously established pathogenic variant regardless of nucleotide change; or protein truncating variant (nonsense, frameshift, canonical +/−1 or 2 splice sites, initiation codon) in a gene where loss of function (LOF) is a known mechanism of disease; and 3) the variant is *de novo* (both maternity and paternity confirmed) in the proband with a negative family history or is inherited from the parents. If the parents are not available for the confirmation of *de novo* or compound heterozygous status of pathogenic variants identified in the proband, the variant would be downgraded and classified as LP. Variants classified as VUS (variant of unknown significance) was defined as the evidence of pathogenicity for the variant is insufficient, or the related disease does not exactly match the patient’s clinical phenotype.

The cohort records the patient’s genetic testing conclusions and corresponding clinical phenotypes. Among them, the phenotypes come from the patient’s clinical records and would be updated during the genetic consultation process. The conclusion of each genetic testing is completed in the following four steps: 1) A junior and a senior genetic counsellors would jointly write conduct the preliminary testing conclusion; 2) The lab director would review the conclusion and issue the report; 3) When necessary, the lab director would discuss the genetic testing conclusions with the patient ‘s clinician, and update the report; 4) For some patients, an updated conclusion would be issued according to the follow-up performance.

### 3. Framework for ConsistencyScore in assisting disease-causing gene prioritization

For simplicity, we named the model generation cohort as CCGT (Children’s Clinical Genetic Testing) Database. This study aims to use ConsistencyScore as the predictor to prioritize P/LP genes from all candidate genes (genes with variants that were identified by NGS and passed the basic filtering including variant QC, target region of sequencing and population allele frequency). The framework included two steps: 1) calculate the cGPS matrix based on clinical genetic reports from CCGT database and public reported databases; and 2) calculate the ConsistencyScore based on the cGPS, the genotype, and the phenotype data of each patient.

#### 3.1 The calculation of cGPS matrix

The cGPS(clinical Gene’s Preferential Synopsis) is a knowledge-based informative matrix which quantifies the gene’s contribution to each clinical category (HPO term or HPO root terms or predefined clinical categories), designed as follows:

Assuming *M* samples from *K* clinical categories (HPO terms or HPO root terms or predefined clinical categories), with *N* genes that were known to cause diseases.

Firstly, for each sample *m*,

a. *S*_*k*_ represents the observation of each clinical category *k*, which is a binary value with value 1 indicating the existence and otherwise 0.
b. *R*_*n*_ is the genetic diagnose level of gene *n*, if gene is diagnosed as P/LP, the value is 1. Specially, for each patient, the original genetic diagnose label at variant-level was directly assigned to gene-level.

Secondly, we applied Naïve Bayes to define cGPS_*k,n*_ as follows, which represents the gene *n*’s ability to cause the observation of a specific affected category *k* if it is the disease-causing gene:

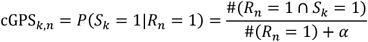

where *#()* represents the set size, e.g #(*R*_*n*_ = 1) represents the number of patients that were diagnosed by gene *n. α* is set as 0.5 to distinguish the condition for the same proportion and more observations will have higher score (e.g 2/(2+0.5) > 1/(1+0.5)). Each cGPS score will provide a p-value representing the confidence of significance by odds ratio between 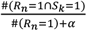 and 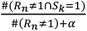.

The purpose of the following steps was to assist prioritization of disease-causing genes. Considering that some genes were never reported as P/LP in the score-generation dataset that would cause the cGPS matrix to zero, false negative results were obtained in the testing dataset for future application. Thus, we added the relationship between genes and clinical synopsis recorded in OMIM, which was collected and curated in two public databases, GeneOrganizer[27] and HPO. We only included “confident” recordings between genes and clinical categories.

For each gene *n*, the modified cGPS_*k,n*_ score for each recorded clinical category *kk* was calculated as follows:

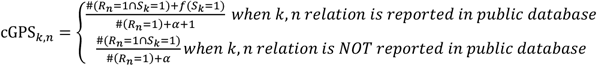

Where 1 in the denominator represents one “fake” observation of *R*_*n*_ = 1. *f*(*S*_*k*_ = 1) in the numerator represents “fake” observation of *R*_*n*_ = 1 ∩ *S*_*k*_ = 1.

#### 3.2 The calculation of ConsistencyScore

For each sample *m* and *K* clinical categories (HPO terms or HPO root terms or predefined clinical categories), for each gene *n*, we defined:

a. *PathogenicityScore* describes the variant or gene’s potential ability to cause disease. For each variant *v*, the PathogenicityScore_*v*_ is assigned by the REVEL [9] score. For variants with PTV (protein truncating variants, VEP annotated as stop_gained, frameshift_variant, splice_acceptor_variant, splice_donor_variant), the PathogenicityScore_*vv*_ is set at 1. For variants annotated with splice_region_variant, the PathogenicityScore_*v*_ is set at 0.5. In sample *m* the PathogenicityScore_*m,v*_ is:

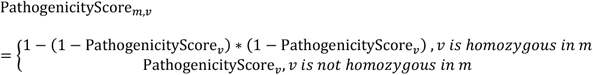 At gene level, the score is calculated as follows:

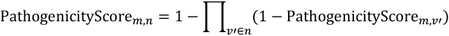 Where *v*′ represents the possible variants in gene *n*.
b. *PhenotypeScore* describes gene’s average phenotype matching effect with no consideration of the pathogenicity. For gene *n* in sample *m*, we averaged the cGPS to this sample’s observed affected clinical category (*S*_*m,k*_ = 1):

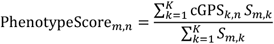
c. *ConsistencyScore* describes the consistency between gene *n*’s predicted affected clinical category to the sample’s observed affected clinical category, defined as:

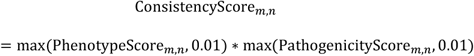

We set the minimum *PathogenicityScore* and *PhenotypeScore* as 0.01 to avoid zero in *ConsistencyScore* that could not be distinguished for ranking. Higher *ConsistencyScore* value represents better consistency between the predicted affected clinical category to the observed ones and higher possibility that gene *n* could be the disease-causing for sample *m*. Considering that in real practice we need to find the disease-causing variants from the disease-causing gene, for variants from the same gene, variant-level PathogenicityScore_*m,v*_ could be used for ranking inside of each gene *m*. In this study, the evaluation and comparison are all at gene-level.

#### 3.3 Evaluation of ConsistencyScore in disease-causing gene’s priorization

Firstly, we randomly chose 70% (14,406) of the samples as the training score-generation dataset to calculate cGPS matrix and applied them to the remaining (6,174) samples as the testing dataset.

Secondly, for each sample *m* and gene *n*, the prediction task is to find out disease-causing sample-gene pairs where *R*_*m,n*_ = 1. Meanwhile, we could obtain PathogenicityScore_*m,n*_, PhenotypeScore_*m,n*_ and ConsistencyScore_*m,n*_ for each sample-gene pair, which we will use as the prediction score. As three strategies were used to calculate cGPS matrix score, here the ConsistencyScore_*m,n*_ were separately calculated based on cGPS matrix score from different strategies and evaluated. Here, one evaluation strategy is to merge all possible sample-genes pairs and to find out whether the higher prediction score could extract out P/LP-diagnosed sample-gene pairs, i.e the disease-causing gene for each sample. Besides, for each sample *m* containing at least one gene with *R*_*m,n*_ = 1, all candidate genes were ranked according to their original prediction scores, and the gene’s rank within each sample were treated as the new prediction score to evaluation the performance. ROC and prevision-recall curves were used to evaluate the performance.

Finally, we applied the prediction framework in a totally independent cohort with 2,323 patients starting from Jan. 2020 to March. 2020 (not covered in the aforementioned model-generation cohort). CGPS matrix were recalculated by using all patient samples. In addition, these 2,323 patient samples went through the normal genetic testing procedure to give report category labels manually for each variant. In total 296 patients had manually reviewed P/LP-diagnosed genes. For each diagnosed patient sample, we ranked the ConsistencyScore (calculated by using predefined clinical categories) in descending order and obtained the ranking value of the final diagnosed disease-causing gene. For each patient sample with multiple P/LP-diagnosted genes, we use the minimum ranking value. Then, for all 296 patient samples, we could summarize the percentage of samples that has the final disease-causing gene with ranking value within top N, where N could be 1,2,3…10.

#### 3.4 Comparison for other tools in disease-causing gene’s priorization

For PathogenicityScore, PheotypeScore, ConsistencyScore and PhenoPro the calculation was based on candidate variant list after the filtration step detailed described above. For the usage of Exomiser, we downloaded source code from https://data.monarchinitiative.org/exomiser/ with version 7.2.1. For the usage of Xrare, we downloaded docker image named xrare-pub-2015.docker.tar.gz and followed instructions from https://web.stanford.edu/~xm24/Xrare/. Considering that the algorithms contain their own filtration step, for Exomiser (Phenix, hiPHIVE) and Xrare, we ran the algorithms by directly input the original VCF file and HPO terms with default parameters. Similarly, we summarized the rank for the causal gene in each candidate list produced by each algorithm for evaluation.

#### 3.5 Application of ConsistencyScore in MGHA corpus

In MGHA corpus, no original genotype files in VCF format could be obtained. Thus, we directly calculate ConsistencyScore (with cGPS matrix score from pre-defined clinical categories) for each reported gene in each sample. For each reported gene, we obtained the distribution for P/LP-reported and not P/LP-reported ConsistencyScore from CCGT dataset and calculated the P/LP and not P/LP probability for the calculated ConsistencyScore in MGHA corpus. We defined predictions with P/LP probability significantly higher than not P/LP probability as a P/LP prediction (OR>2). For the HPO terms recorded, we manually check their status in the current HPO database and replace ones with “obsolete” condition to the replaced terms and “alt_id” ones to the main term. i.e replace “HP:0002281” to “HP:0002282”, “HP:0005549” to “HP:0001875”, “HP:0008012” to “HP:0000545”, “HP:0008538” to “HP:0000407”, “HP:0006996” to “HP:0006989”.

### 4. Statistical analysis

All statistical analysis was performed by R version 3.6.1. Student’s t-test was used for pairwise numeric vector comparison, realized by t.test() in R. The confidence interval (CI) for the percentage of cases were performed by bootstrap strategy with 1000 times. If the original sample size is smaller than 10, no CI is provided. The comparison for prediction performance was realized by the proportion test for the ranked-first percentage, realized by prop.test() in R. The CI and comparison between AUC values (ROC) were used by R package pROC by setting method= ‘delong’. For the precision-recall curve, the CI was obtained by bootstrap strategy with 1000 times and the comparison between AUC values were tested by t.test() for the bootstrapped result vector.

## Data Availability

Dr. Wenhao Zhou, National Children's Medical Center, Children's Hospital of Fudan University, Shanghai, China, had full access to all the data in the study and takes responsibility for the integrity of the data and the accuracy of the data analysis. The cGPS matrix score generated during the current study are available from the corresponding author upon reasonable request.

## Declarations

### Ethics approval and consent to participate

The collection of human samples was approved by the ethics committees of Children’s Hospital of Fudan University and informed consent was taken from all individual participants. This study was performed in line with the principles of the Declaration of Helsinki.

### Code availability

The code to calculate cGPS and ConsistencyScore during the current study are available from the corresponding author upon reasonable request.

### Authors’ contributions

Yulan Lu and Wenhao Zhou conceived the project. Xinran Dong, Bingbing Wu, Qi Ni, Yaqiong Wang, and Bo Liu performed computational analyses. Huijun Wang, Lin Yang, Xiang Chen collected the human samples. Xinran Dong and Bingbing Wu wrote the manuscript. All authors have read and approved the manuscript.

## Acknowledgements

We thank all doctors and nurses in our hospital for their patient care and data collection. We are very grateful to the patient families for their trust in our lab.

