## Supplementary material for "An automatic diagnostic system for pediatric genetic disorders by linking genotype and phenotype information": supp-1.docx

### Supplementary Figures

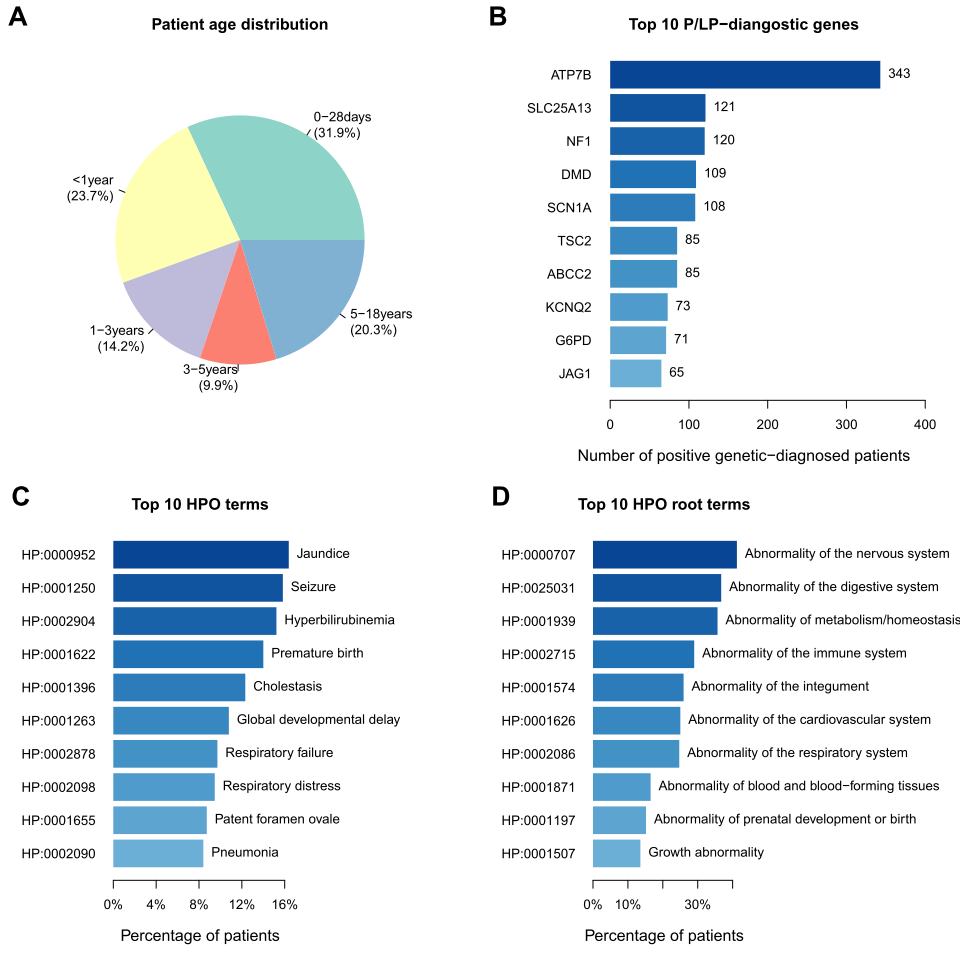

**Supplementary Figure1. Basic statistics for patients included in the model-generation cohort.** (A) Age distribution for all patients. (B) Top 10 P/LP-diagnosed genes. The number represent the number of positive genetic-diagnosed patients. (C) Top 10 HPO terms. (D) Top 10 HPO root terms (the first level child nodes under HP:0000118: Phenotype abnormality).

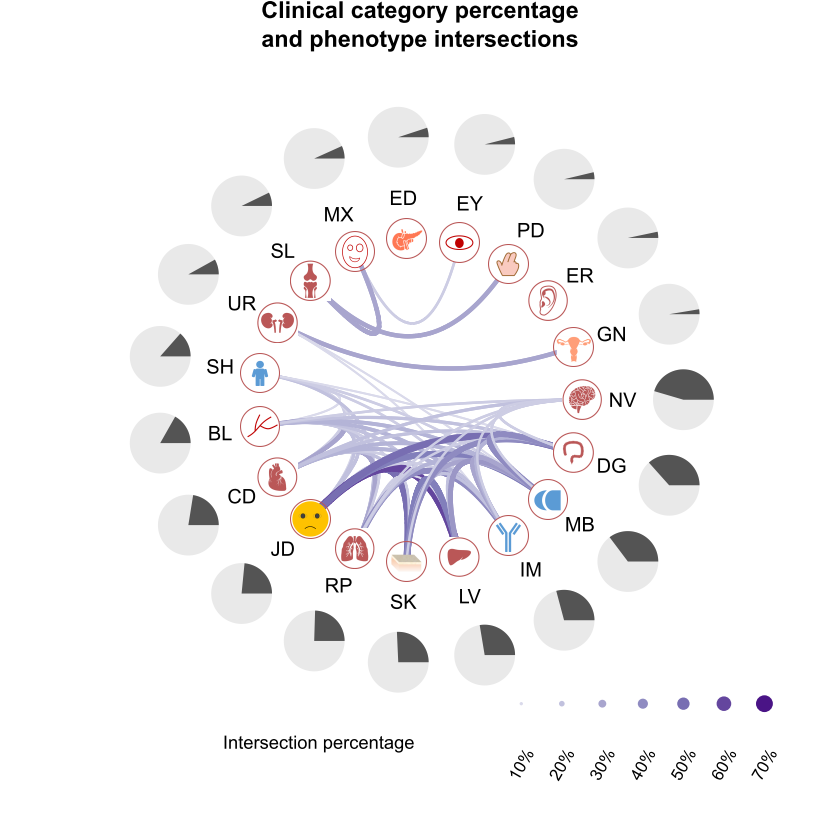

**Supplementary Figure2.** Percentage of patients affected by each pre-defined clinical category and phenotype intersections. Category percentages were shown in separated pie chart, while the intersection between categories higher than 10% were shown. The line color and width were proportion to the intersection percentage.

Abbreviation:

NV: Nervous system; DG: Digestive; MB: Metabolism; IM: Immune; LV: Liver; SK: Skin; PR: Respiratory system; JD: Jaundice; CD: Cardiovascular system; BL: Blood; UR: Urinary; SL: Skeletal; MX: Maxillofacial deformity; ED: Endocrine; SH: Short stature; EY: Eye; PD: Polydactyly/Abnormal of limbs; ER: Ear; GN: Genital system;

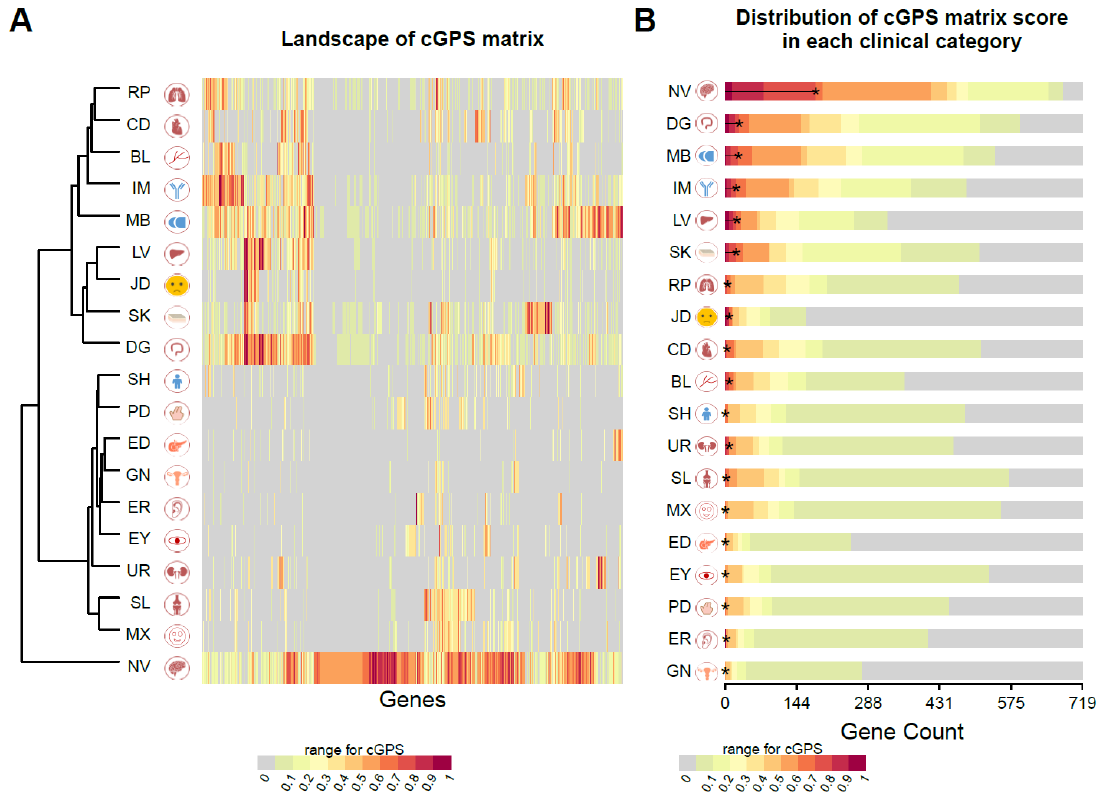

**Supplementary Figure3. cGPS (clinical Gene’s Preferential Synopsis) matrix describes the diagnostic contribution of genes to phenotypes grouped as clinical categories.** **(A)** Heatmap for the cGPS matrix scores of different genes in different clinical categories. The dendrogram shows category similarity measured by the euclidian distance and using ward. D as the hierarchical clustering strategy. **(B)** Distribution of cGPS matrix score in each clinical category (score range shown in color). Genes were ranked by cGPS, and 0.7 cutoff was marked by the “*”. In this figure, genes with maximum cGPS score higher than 0.5 across all clinical categories were shown.

Abbreviation:

NV: Nervous system; DG: Digestive; MB: Metabolism; IM: Immune; LV: Liver; SK: Skin; PR: Respiratory system; JD: Jaundice; CD: Cardiovascular system; BL: Blood; UR: Urinary; SL: Skeletal; MX: Maxillofacial deformity; ED: Endocrine; SH: Short stature; EY: Eye; PD: Polydactyly/Abnormal of limbs; ER: Ear; GN: Genital system;

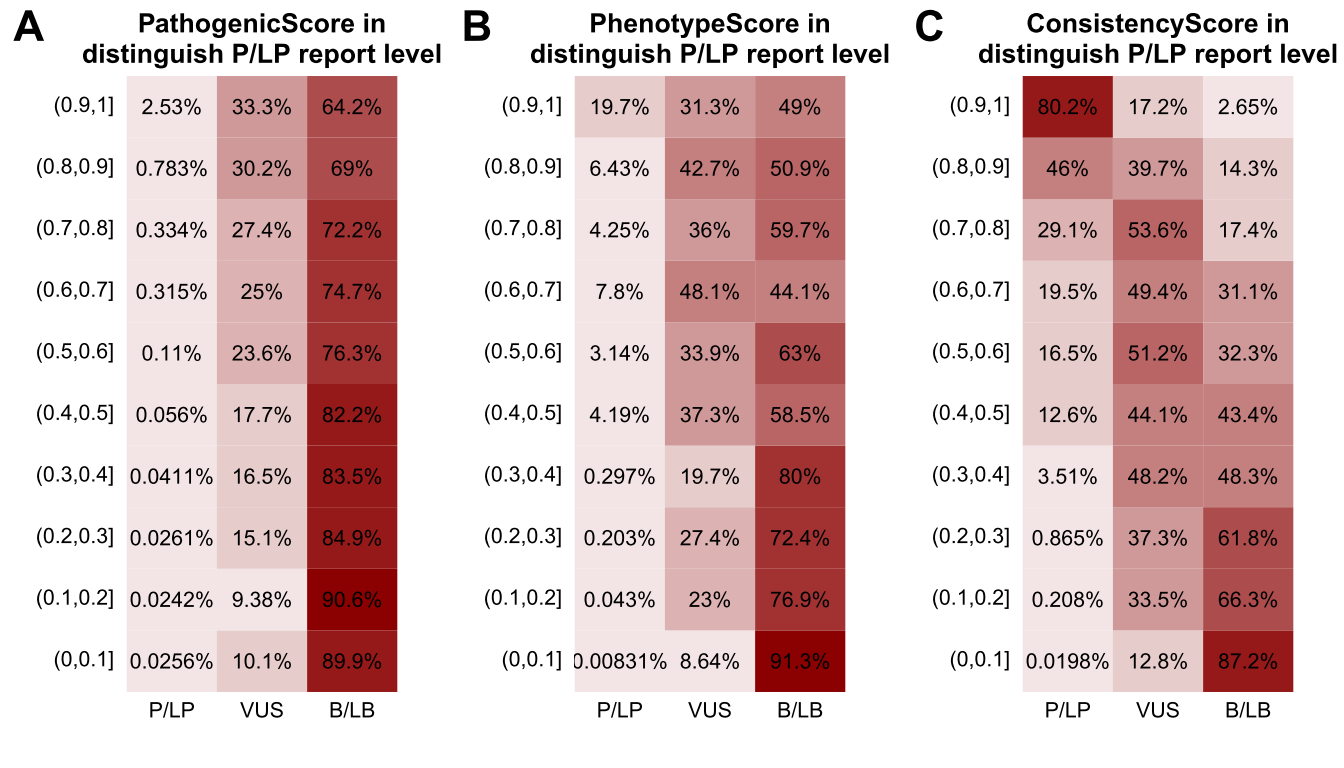

**Supplementary Figure4. Percentage of case-gene pairs reported as P/LP, VUS and B/LB with PathogenicityScore (A), PhenotypeScore (B) and ConsistencyScore (C) within each value interval.** For example, if focus on ConsistencyScore higher than 0.9, 80.2% of the case-gene pairs were reported as P/LP. P: positive, LP: likely positive, B: benign, LB: likely benign.

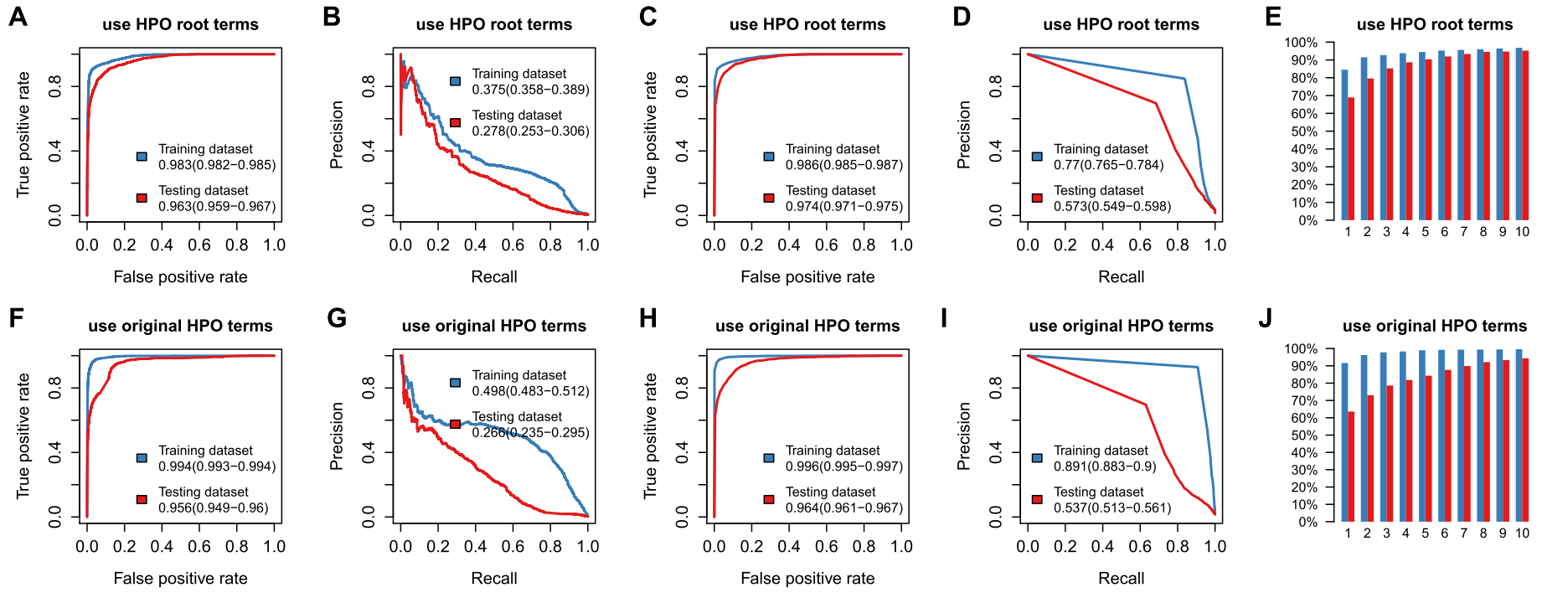

**Supplementary Figure5. Performance of ConsistencyScore based on cGPS matrix score using HPO root terms (A-E) and original HPO terms (F-J).** (A, F) The performance of using the ConsistencyScore to predict diagnostic sample-gene pairs in the training and testing dataset, evaluated by the ROC curves. (B, G) The performance of using the ConsistencyScore to predict diagnostic sample-gene pairs in the training and testing dataset, evaluated by the precision-recall curves. (C, H) The performance of using gene’s rank ordered by ConsistencyScore to predict diagnostic sample-gene pairs in the training and testing dataset, evaluated by the ROC curves. (D, I) The performance of using gene’s rank ordered by ConsistencyScore to predict diagnostic sample-gene pairs in the training and testing dataset, evaluated by the precision-recall curves. (E,J) The percentage of cases with final diagnostic-genes within top candidate number (1,2…10) in training dataset (blue) and testing dataset (red), either using HPO root terms (E) or using the original HPO terms (J).

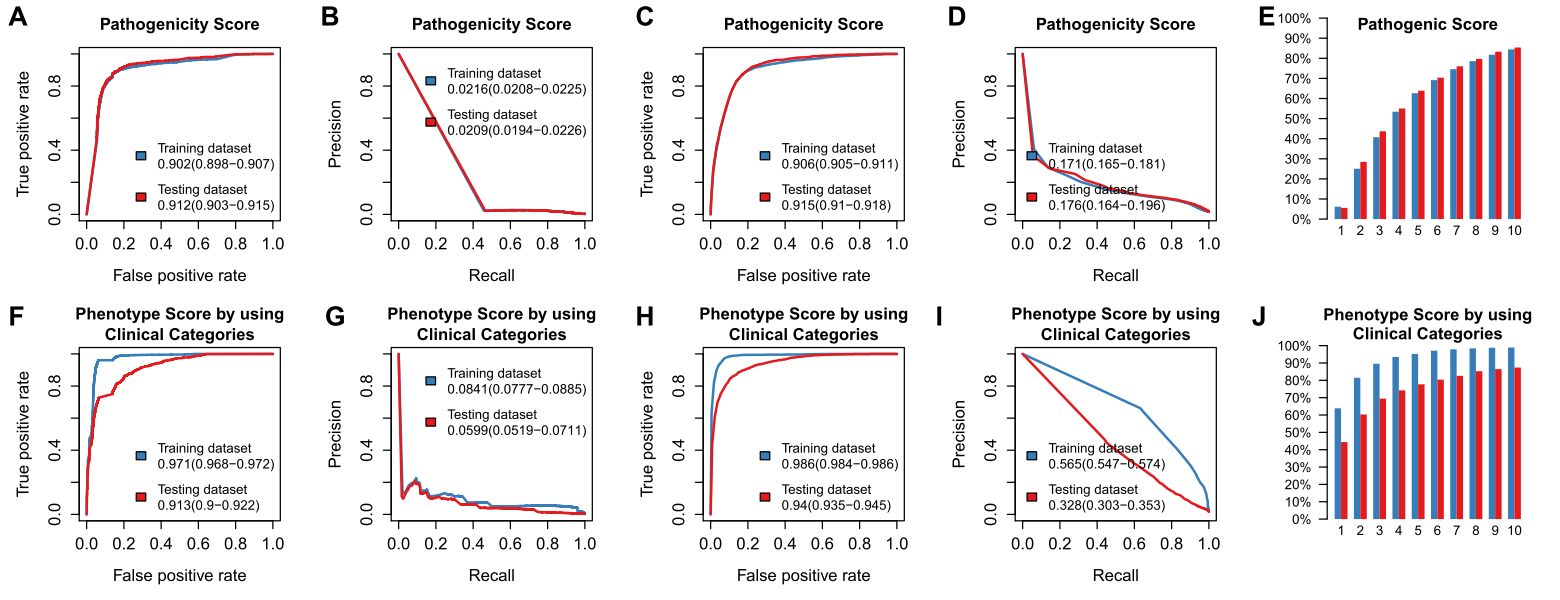

**Supplementary Figure6. Performance of PathogenicityScore (A-E) and PhenotypeScore (F-J) based on cGPS matrix score using predefined clinical categories.** (A, F) The performance of PathogenicityScore and PhenotypeScore in ROC curves to predict diagnostic sample-gene pairs in training and testing dataset. (B, G) The performance of PathogenicityScore and PhenotypeScore in precision-recall curves to predict diagnostic sample-gene pairs in training and testing dataset. (C, H) The performance of gene’s rank ordered by PathogenicityScore and PhenotypeScore in ROC curves to predict diagnostic sample-gene pairs in training and testing dataset. (D, I) The performance of gene’s rank ordered by PathogenicityScore and PhenotypeScore in precision-recall curves to predict diagnostic sample-gene pairs in training and testing dataset. (E,J) The percentage of cases with final diagnostic-genes within top candidate number (1,2…10) in training dataset (blue) and testing dataset (red).

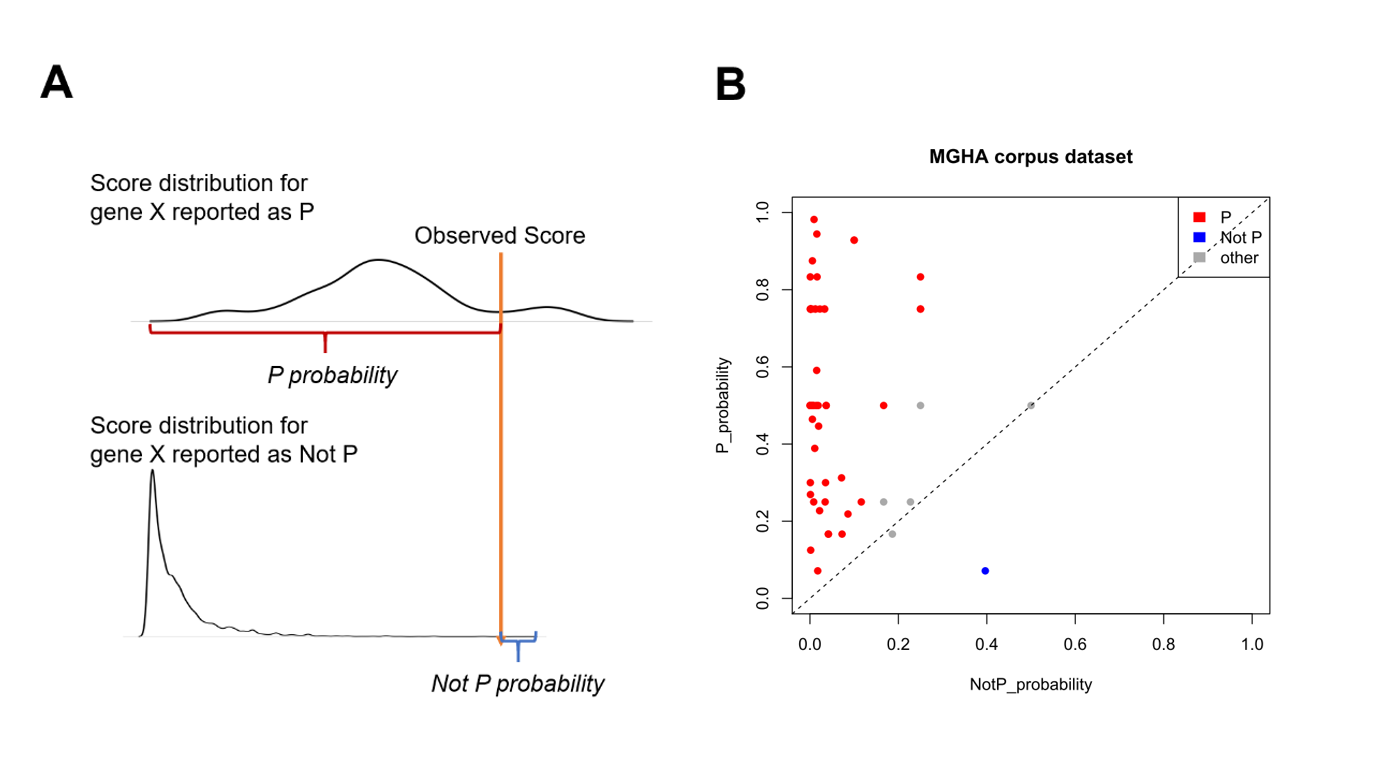

**Supplementary Figure7. Performance of ConsistencyScore in MGHA corpus.** (A) The evaluation strategy of ConsistencyScore in predicting P/LP (short write as P) genes. For each reported gene, we obtained the distribution for P/LP-reported and not P/LP-reported ConsistencyScore from CCGT dataset and calculated the P/LP and not P/LP probability for the observed score in MGHA corpus based on the distribution. (B) Performance of ConsistencyScore in P probability (Y-axis) and Not P probability (X-axis) in 59 variants of 47 MGHA samples. We defined predictions with P/LP probability significantly higher than not P/LP probability (OR>2) as a P/LP prediction (marked as P). Predictions with P/LP probability significantly lower than not P/LP probability (OR<0.5) as not P/LP prediction (marked as Not P). Others are marked as other.

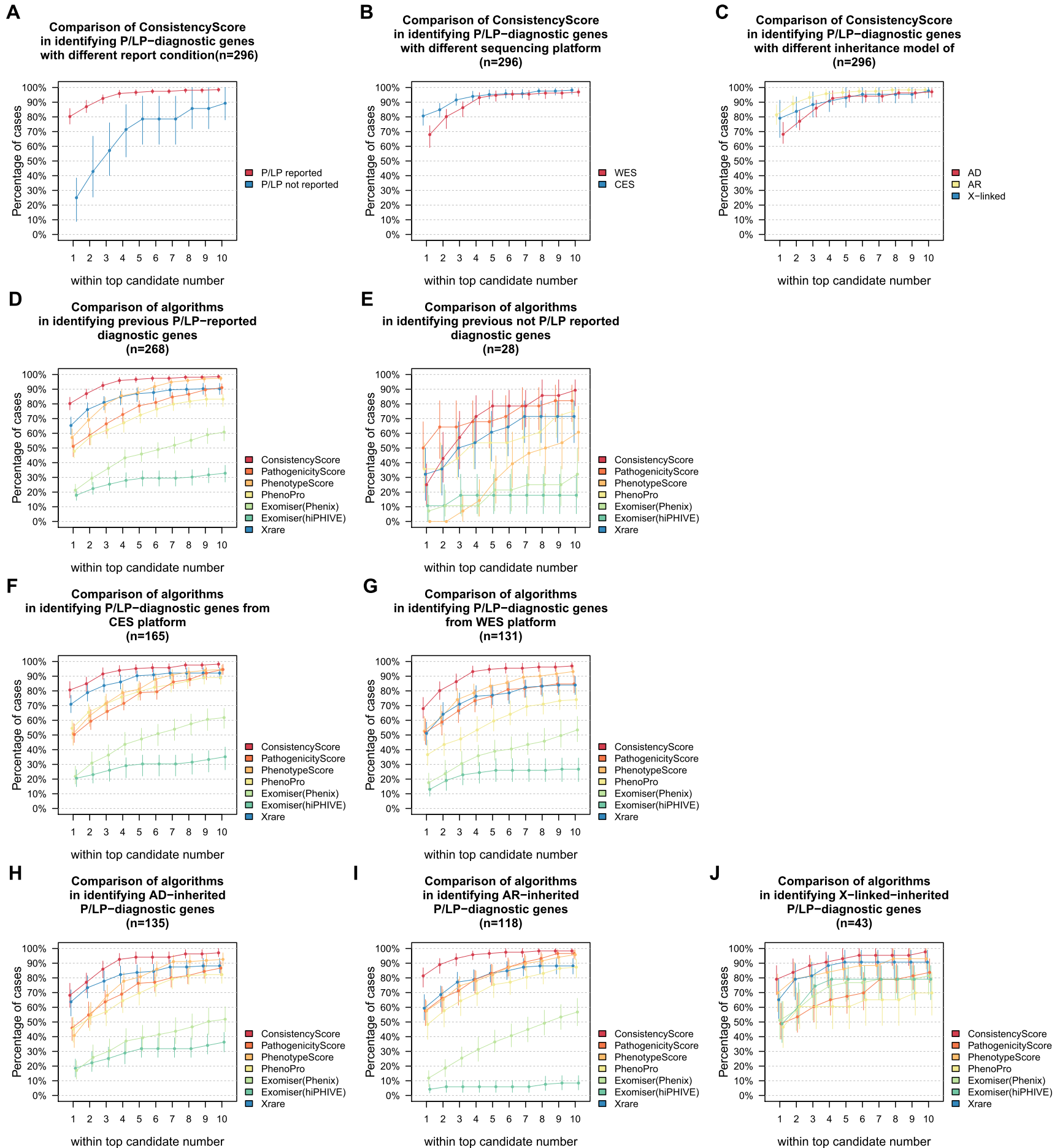

**Supplementary Figure8. Performance evaluation in priorization disease-causing genes from 296 P/LP-diagnosed patients in different scenarios.** (A-C) Performance of ConsistencyScore for genes with model-generation cohort P/LP reported or not (A), sequenced by CES or WES platform (B) and genes with different inheritance model (C). (D-J) Performance comparison for different tools in samples with different scenarios. (D) P/LP reported (E) not P/LP reported (F) sequenced by CES platform (G) sequenced by WES platform (H) genes with AD-inherited model (I) AR-inherited model (J) X-linked-inherited model. For each sub-plot, the cumulative percentage of samples with final diagnostic-genes within top candidate number (1,2…10) produced were shown. For P/LP reported, cGPS matrix is mainly calculated by information from real P/LP observations. Otherwise, extended cGPS matrix is inferred from public databases. For each sub-figure, 95% CI bar were shown.

### Supplementary Tables

**Supplementary Tables 1**. Detailed description of 19 clinical categories and related HPO terms.

|  | Test Type | |
| --- | --- | --- |
|  | **WES**  **(whole exome sequencing)** | **CES**  **(clinical exome sequencing)** |
| Exome capture Kit | Agilent Sureselect  All Exons Human V5 Kit | Agilent ClearSeq  Inherited Disease Kit |
| Sequencing platform | Illumina HiSeq 2000/X10, 150PE | Illumina HiSeq 2000/X10, 150PE |
| Average reads number (M) | 67.62 | 26.52 |
| Average reads mapping rate | 99.85% | 99.83% |
| Mean coverage sequencing depth on official target | 126.14 | 230.4 |

**Supplementary Tables 2**. Detailed description of 19 clinical categories and related HPO terms.

| **HPO_Terms** | **HPO_Name** | **Clinical_category** | **Abbreviation** |
| --- | --- | --- | --- |
| HP:0000952 | Jaundice | Jaundice | JD |
| HP:0001396 | Cholestasis | Jaundice | JD |
| HP:0001406 | Intrahepatic cholestasis | Jaundice | JD |
| HP:0002904 | Hyperbilirubinemia | Jaundice | JD |
| HP:0003265 | Neonatal hyperbilirubinemia | Jaundice | JD |
| HP:0000707 | Abnormality of the nervous system | Nervous system | NV |
| HP:0000717 | Autism | Nervous system | NV |
| HP:0000729 | Autistic behavior | Nervous system | NV |
| HP:0000750 | Delayed speech and language development | Nervous system | NV |
| HP:0001249 | Intellectual disability | Nervous system | NV |
| HP:0001250 | Seizure | Nervous system | NV |
| HP:0001263 | Global developmental delay | Nervous system | NV |
| HP:0001268 | Mental deterioration | Nervous system | NV |
| HP:0001270 | Motor delay | Nervous system | NV |
| HP:0001288 | Gait disturbance | Nervous system | NV |
| HP:0002141 | Gait imbalance | Nervous system | NV |
| HP:0001298 | Encephalopathy | Nervous system | NV |
| HP:0002353 | EEG abnormality | Nervous system | NV |
| HP:0002373 | Febrile seizure (within the age range of 3 months to 6 years) | Nervous system | NV |
| HP:0002376 | Developmental regression | Nervous system | NV |
| HP:0006801 | Hyperactive deep tendon reflexes | Nervous system | NV |
| HP:0009830 | Peripheral neuropathy | Nervous system | NV |
| HP:0011097 | Epileptic spasm | Nervous system | NV |
| HP:0012469 | Infantile spasms | Nervous system | NV |
| HP:0012758 | Neurodevelopmental delay | Nervous system | NV |
| HP:0002333 | Motor deterioration | Nervous system | NV |
| HP:0100660 | Dyskinesia | Nervous system | NV |
| HP:0002104 | Apnea | Nervous system | NV |
| HP:0002355 | Difficulty walking | Nervous system | NV |
| HP:0000639 | Nystagmus | Nervous system | NV |
| HP:0000252 | Microcephaly | Nervous system | NV |
| HP:0002059 | Cerebral atrophy | Nervous system | NV |
| HP:0002500 | Abnormality of the cerebral white matter | Nervous system | NV |
| HP:0006872 | Cerebral hypoplasia | Nervous system | NV |
| HP:0001252 | Muscular hypotonia | Nervous system | NV |
| HP:0001276 | Hypertonia | Nervous system | NV |
| HP:0001290 | Generalized hypotonia | Nervous system | NV |
| HP:0001319 | Neonatal hypotonia | Nervous system | NV |
| HP:0001324 | Muscle weakness | Nervous system | NV |
| HP:0001332 | Dystonia | Nervous system | NV |
| HP:0002141 | Gait imbalance | Nervous system | NV |
| HP:0002317 | Unsteady gait | Nervous system | NV |
| HP:0002333 | Motor deterioration | Nervous system | NV |
| HP:0003198 | Myopathy | Nervous system | NV |
| HP:0003236 | Elevated serum creatine kinase | Nervous system | NV |
| HP:0003560 | Muscular dystrophy | Nervous system | NV |
| HP:0040081 | Abnormal circulating creatine kinase concentration | Nervous system | NV |
| HP:0100021 | Cerebral palsy | Nervous system | NV |
| HP:0002355 | Difficulty walking | Nervous system | NV |
| HP:0100660 | Dyskinesia | Nervous system | NV |
| HP:0003011 | Abnormality of the musculature | Nervous system | NV |
| HP:0011968 | Feeding difficulties | Nervous system | NV |
| HP:0100697 | Neurofibrosarcoma | Nervous system | NV |
| HP:0010614 | Fibroma | Nervous system | NV |
| HP:0000486 | Strabismus | Eye | EY |
| HP:0000518 | Cataract | Eye | EY |
| HP:0000478 | Abnormality of the eye | Eye | EY |
| HP:0000639 | Nystagmus | Eye | EY |
| HP:0000598 | Abnormality of the ear | Ear | ER |
| HP:0000365 | Hearing impairment | Ear | ER |
| HP:0000356 | Abnormality of the outer ear | Ear | ER |
| HP:0000359 | Abnormality of the inner ear | Ear | ER |
| HP:0000370 | Abnormality of the middle ear | Ear | ER |
| HP:0000152 | Abnormality of head or neck | maxillofacial deformity | MX |
| HP:0000218 | High palate | maxillofacial deformity | MX |
| HP:0000347 | Micrognathia | maxillofacial deformity | MX |
| HP:0001601 | Laryngomalacia | maxillofacial deformity | MX |
| HP:0000252 | Microcephaly | maxillofacial deformity | MX |
| HP:0000175 | Cleft palate | maxillofacial deformity | MX |
| HP:0002086 | Abnormality of the respiratory system | Respiratory system | RP |
| HP:0002092 | Pulmonary arterial hypertension | Respiratory system | RP |
| HP:0002098 | Respiratory distress | Respiratory system | RP |
| HP:0002878 | Respiratory failure | Respiratory system | RP |
| HP:0012387 | Bronchitis | Respiratory system | RP |
| HP:0002104 | Apnea | Respiratory system | RP |
| HP:0002090 | Pneumonia | Respiratory system | RP |
| HP:0012252 | Abnormal respiratory system morphology | Respiratory system | RP |
| HP:0001626 | Abnormality of the cardiovascular system | Cardiovascular system | CD |
| HP:0011025 | Abnormal cardiovascular system physiology | Cardiovascular system | CD |
| HP:0001631 | Atrial septal defect | Cardiovascular system | CD |
| HP:0001643 | Patent ductus arteriosus | Cardiovascular system | CD |
| HP:0001655 | Patent foramen ovale | Cardiovascular system | CD |
| HP:0001627 | Abnormal heart morphology | Cardiovascular system | CD |
| HP:0002631 | Ascending aortic aneurysm | Cardiovascular system | CD |
| HP:0001928 | Abnormality of coagulation | Blood | BL |
| HP:0001871 | Abnormality of blood and blood-forming tissues | Blood | BL |
| HP:0001903 | Anemia | Blood | BL |
| HP:0001881 | Abnormal leukocyte morphology | Blood | BL |
| HP:0001873 | Thrombocytopenia | Blood | BL |
| HP:0001872 | Abnormal thrombocyte morphology | Blood | BL |
| HP:0001942 | Metabolic acidosis | Metabolism | MB |
| HP:0001943 | Hypoglycemia | Metabolism | MB |
| HP:0001987 | Hyperammonemia | Metabolism | MB |
| HP:0001998 | Neonatal hypoglycemia | Metabolism | MB |
| HP:0002901 | Hypocalcemia | Metabolism | MB |
| HP:0004395 | Malnutrition | Metabolism | MB |
| HP:0001939 | Abnormality of metabolism/homeostasis | Metabolism | MB |
| HP:0002013 | Vomiting | Metabolism | MB |
| HP:0010837 | Decreased serum ceruloplasmin | Metabolism | MB |
| HP:0000821 | Hypothyroidism | Endocrine | ED |
| HP:0000818 | Abnormality of the endocrine system | Endocrine | ED |
| HP:0001945 | Fever | Immune | IM |
| HP:0001954 | Recurrent fever | Immune | IM |
| HP:0002721 | Immunodeficiency | Immune | IM |
| HP:0100806 | Sepsis | Immune | IM |
| HP:0002715 | Abnormality of the immune system | Immune | IM |
| HP:0002090 | Pneumonia | Immune | IM |
| HP:0002014 | Diarrhea | Immune | IM |
| HP:0025031 | Abnormality of the digestive system | Digestive | DG |
| HP:0004395 | Malnutrition | Digestive | DG |
| HP:0002014 | Diarrhea | Digestive | DG |
| HP:0002013 | Vomiting | Digestive | DG |
| HP:0025032 | Abnormality of digestive system physiology | Digestive | DG |
| HP:0025033 | Abnormality of digestive system morphology | Digestive | DG |
| HP:0001392 | Abnormality of the liver | Liver | LV |
| HP:0001410 | Decreased liver function | Liver | LV |
| HP:0002240 | Hepatomegaly | Liver | LV |
| HP:0002910 | Elevated hepatic transaminase | Liver | LV |
| HP:0006568 | Increased hepatic glycogen content | Liver | LV |
| HP:0001433 | Hepatosplenomegaly | Liver | LV |
| HP:0001394 | Cirrhosis | Liver | LV |
| HP:0001399 | Hepatic failure | Liver | LV |
| HP:0100839 | Hepatic agenesis | Liver | LV |
| HP:0030146 | Abnormal liver parenchyma morphology | Liver | LV |
| HP:0000119 | Abnormality of the genitourinary system | Urinary | UR |
| HP:0012622 | Chronic kidney disease | Urinary | UR |
| HP:0011277 | Abnormality of the urinary system physiology | Urinary | UR |
| HP:0000809 | Urinary tract atresia | Urinary | UR |
| HP:0010936 | Abnormality of the lower urinary tract | Urinary | UR |
| HP:0010935 | Abnormality of the upper urinary tract | Urinary | UR |
| HP:0000924 | Abnormality of the skeletal system | Skeletal | SL |
| HP:0002650 | Scoliosis | Skeletal | SL |
| HP:0001601 | Laryngomalacia | Skeletal | SL |
| HP:0000924 | Abnormality of the skeletal system | Skeletal | SL |
| HP:0010442 | Polydactyly | Polydactyly/Abnormal of limbs | PD |
| HP:0040064 | Abnormality of limbs | Polydactyly/Abnormal of limbs | PD |
| HP:0001510 | Growth delay | Short stature | SH |
| HP:0004322 | Short stature | Short stature | SH |
| HP:0001507 | Growth abnormality | Short stature | SH |
| HP:0000953 | Hyperpigmentation of the skin | Skin | SK |
| HP:0000957 | Cafe-au-lait spot | Skin | SK |
| HP:0000964 | Eczema | Skin | SK |
| HP:0001067 | Neurofibromas | Skin | SK |
| HP:0001574 | Abnormality of the integument | Skin | SK |
| HP:0000078 | Abnormality of the genital system | genital system | GN |

**Supplementary Tables 3**. Performance of the ConsistencyScore in the MGHA cohort.
